# Machine-Learning based Prediction Models for Healthcare Outcomes in Patients Participating in Cardiac Rehabilitation: A Systematic Review

**DOI:** 10.1101/2024.07.09.24310007

**Authors:** Xiarepati Tieliwaerdi, Kathryn Manalo, Abulikemu Abuduweili, Sana Khan, Edmund Appiah-kubi, Andrew Oehler

**Affiliations:** Department of Medicine, Allegheny Health Network, Pittsburgh, Pennsylvania; Institute of Robotics, Carnegie Mellon University, Pittsburgh, Pennsylvania; Allegheny Health Network Cardiovascular Institute, Allegheny Health Network, Pittsburgh, Pennsylvania

## Abstract

**Purpose:** CR has been proven to reduce mortality and morbidity in patients with CVD. ML techniques are increasingly used to predict healthcare outcomes in various fields of medicine including CR. This systemic review aims to perform critical appraisal of existing ML based prognosis predictive model within CR and identify key research gaps in this area.

**Review methods:** A systematic literature search was conducted in Scopus, PubMed, Web of Science and Google Scholar from the inception of each database to 28th January 2024. The data extracted included clinical features, predicted outcomes, model development and validation as well as model performance metrics. Included studies underwent quality assessments using the IJMEDI.

**Summary:** 22 ML-based clinical models from 7 studies across multiple phases of CR were included. Most models were developed using smaller patient cohorts from 41 to 227, with one exception involving 2280 patients. The prediction objectives ranged from patient intention to initiate CR to graduate from outpatient CR along with interval physiological and psychological response to CR. The best-performing ML models reported AUC between 0.82 and 0.91, sensitivity from 0.77 to 0.95, indicating good prediction capabilities. However, none of them underwent calibration or external validation. Most studies raised concerns for bias. Readiness of these models for implement into practice is questionable. External validation of existing models and development of new models with robust methodology based on larger populations and targeting diverse clinical overcomes in CR are needed.

## Introduction

Cardiovascular disease (CVD) remains a leading cause of morbidity and mortality globally, causing over 19 million deaths alone in 2020 worldwide^1^. Cardiac rehabilitation (CR) is a comprehensive evidenced-based intervention tailored to patients with CVD conditions like ischemic heart disease, heart failure, myocardial infarction (MI), or those undergoing cardiovascular interventions such as coronary angioplasty or bypass grafting^2–5^. It encompasses exercise training, health behavior modification, patient education, nutritional and psychological counseling, proven to reduce morbidity and mortality, enhance functional capacity, and improve quality of life in CVD patients^6–11^. CR comprises of three phases in general. Phase I occurs post-cardiovascular events during the acute inpatient setting for stabilization and minimizing deconditioning. Phase II begins once the patient is able to be safely discharged from the hospital. A patient can expect to participate in outpatient CR focusing on supervised exercise program and medical education generally up to 12 weeks with 36 sessions in total. After completion, patients enter the phase III as a long-term maintenance period, where patients continue their rehab exercises and lifestyle changes on their own at home or within a community setting^12–14^.

CR is a complex medical entity, where challenges lie in multiple aspects including unsatisfactory patient enrollment and adherence, varying clinical responses and the need for ongoing treatment adjustment to improve ultimate outocmes^15–18^. CR program involves a large amount of multimodal electronic health records (EHRs), which contain a wide range of variables in diverse data formats such as demographic information, providers’ free-text notes, laboratory and imaging findings. These data are often too voluminous and technically unfeasible to process by traditional analytical methods^19^. In addition, traditional statistical methods are built on specific assumptions, including specific error distributions, the additive linear predictor’s parameters, and the proportionality of hazards^20^. However, these assumptions do not always align with the realities encountered in clinical settings. Moreover, traditional methods primarily aim to infer relationships between variables, particularly interactions between a main determinant and individual confounders, they are insufficient in current clinical settings, where there are a growing number of variables and an increasing need for early intervention through outcome prediction^21^. The advent of machine learning (ML) in healthcare offers a potential solution to these challenges. ML detects intricate interactions among millions of complex variables without being programed with hypophysis^22^. It is particularly well-suited for analyzing large datasets with numerous features and high complexities such as EHR. It can analyze various data types and is better at capturing non-linear relationships between variables^20,23^. ML based predictive models have been widely proposed in CVD to aid decision-making processes for clinicians, showing promise in predicting patient specific clinical progression and health care outcomes^24–26^.

Recent literature has seen a proliferation of systematic reviews and meta-analyses targeted at evaluating the performance of ML-based clinical prediction models in different fields of medicine^27–29^. However, to the best of our knowledge, there has yet to be a systematic review of ML predictive models in CR. The primary aim of this review is to evaluate existing literature regarding ML predictive model in the context of CR and provide systemic appraisal of these models from their development to validation. We aim to compare performance metrics, validation processes and appropriateness of algorithm used in CR, as well as identify key research gaps in this area.

## Methods

### Literature Search Strategy

The literature search and related statistical analyses were conducted in accordance with the guidelines of the Preferred Reporting Items for Systematic Reviews and Meta-Analyses (PRISMA) Statement^30^. We comprehensively searched publications from database inception to 28th January 2024 in Google Scholar, PubMed, Web of Science, and Scopus. Keywords used for the search included a combination of terms related to ML and CR. The full search strategy, including the specific combinations of search terms used in each database, was provided in supplementary table 1.

### Inclusion and Exclusion Criteria

In our review, we included studies that employed a ML model to predict clinical progression, health outcomes, or risk stratification in a cohort of adult patients participating in any phase of CR. We did not restrict it by the country of origin or publication source. For clarity, we defined ML as algorithms, such as random forest (RF) analysis, support vector machines (SVM), and neural networks, that are more complex than logistic regression models and capable of making decisions based on data patterns.

Our inclusion criteria were structured using the population, intervention, comparison, outcome, and time (PICOT) approach as follows^31^: 1) Population: Adult patients aged 18 years or older engaging in CR programs; 2) Intervention: Studies used ML models for predicting outcomes in CR; 3) Comparison: Not applicable, due to the lack of a universally accepted prognostic model in CR; 4) Outcome: Studies reporting on clinical progression, health outcomes or risk stratification outcomes; 5) Time: Studies which harnessed features to predict outcomes after any given follow-up period is accepted. In terms of study design, we did not restrict the types of studies included. Retrospective, prospective, cross-sectional, cohort studies, case-control studies and randomized controlled trials were all considered for inclusion.

Exclusion criteria were: 1) Studies not in English; 2) Studies not involving ML-based predictive models: for example, studies reporting novel ML based wearable devices used for monitoring biomarkers such as heart rate or blood pressure were excluded; 3) Studies where predictive models are not developed from patients in CR; 4) Studies focusing on identifying predictors associated with outcomes rather than develop a prognostic model. 5) Reviews, case reports and studies not available in full text were excluded.

### Data Extraction

Duplicate records were initially removed using auto-deduplication function in EndNote 21, followed by a manual check for complete deduplication. The screening of titles and abstracts was then carried out in EndNote 21, adhering to the inclusion and exclusion criteria previously outlined.

A team of four reviewers (XT, KM, SK and EA) assessed articles for eligibility first by screening titles and abstracts to ensure relevance in EndNote; each study was independently assessed by at least two reviewers. Both agreed-upon and conflicting articles were retained for second-round screening based on full-text review. The full-text evaluation was independently carried out by the reviewers (XT and AA). Disagreements were resolved through mutual consensus.

The data from included articles were extracted into tables by two authors (SK and KM) independently. Cross-check was conducted. Variables were extracted and tabulated in Excel 2020, which includes 1) baseline characteristics of studies; 2) features and ML algorithms used for the development of the models; 3) objectives of the prediction models as well as evaluation index for their performance, including area under the receiver operating characteristic curve (AUROC), accuracy, precision, sensitivity and specificity; 4) methods employed for model validation.

### Quality Assessment

The quality of the included studies was evaluated independently by two reviewers (XT and AA) by using IJMEDI checklist^32^. The IJMEDI checklist is developed specifically for use in medical ML studies to distinguish high-quality ML research from studies with mere application of ML methods to medical data. It focuses on six key aspects: problem and data understanding, data preparation, modeling, validation, and deployment, encompassing a total of 30 detailed questions. Responses to these questions are categorized as OK (adequately addressed), mR (moderately addressed with improvement potential), or MR (inadequately addressed). For questions deemed high priority, scores were assigned as 0 for OK, 1 for mR, and 2 for MR. For low-priority questions, The scoring was assigned as 0 for OK, 1 for mR, and 2 for MR. Studies were classified into three quality categories: low (0–19.5 points), medium (20–34.5 points), or high quality (35–50 points).

## Results

From the initial search that yielded 151 records, 81 were excluded following a review of the title and abstract, 60 records were excluded after full-text review. In the end, 7 studies met the inclusion criteria and were included in our systemic review (Figure 1 shows process of screening and study selection)^35–41^.

**Figure 1:**
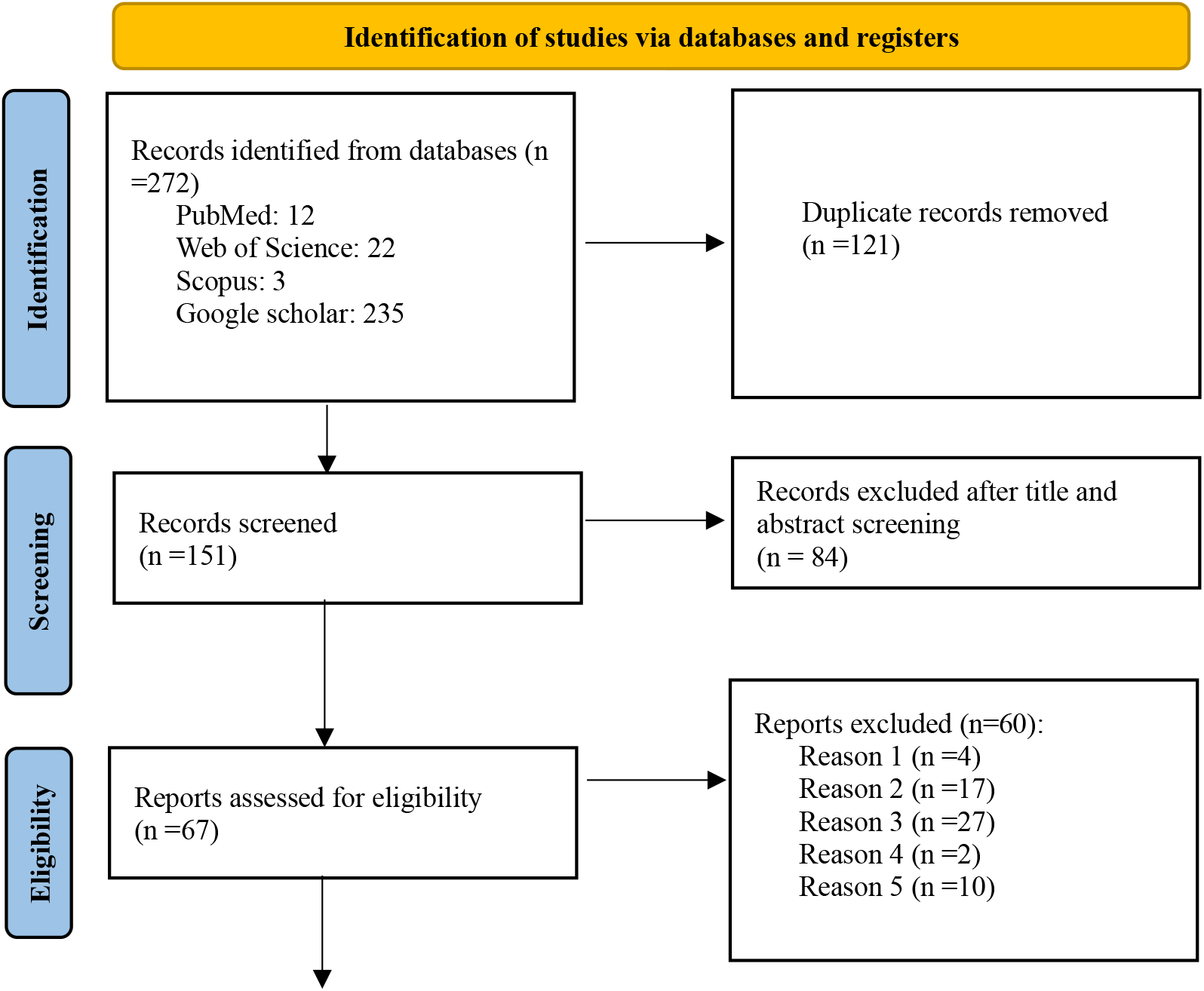
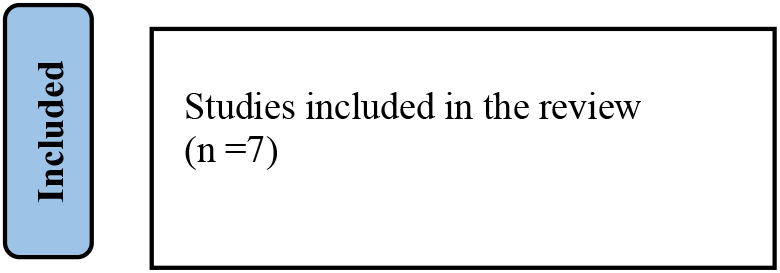
PRISMA flowchart of the review process

### Study characteristics

Table 1 summarizes characteristics of studies using ML prediction model in CR. Seven studies are included, one of which is a multicenter study conducted in Belgium and Ireland^37^. The rest are single-center studies from Australia^35,40^, Belgium^38^, Italy^41^, Malaysia^36^, and Chile^39^. Study designs include five retrospective studies^35–38,41^, one cross-sectional study^40^, and one that combines retrospective and prospective approaches^39^. Most studies target phase II of CR, with one addressing phases II and III^35^ and another focusing solely on phase III^37^. Participant ages were reported in four of the seven studies^35,38,40,41^, with mean ages ranging from 63 to 67.98 years. The percentage of female participants is reported in three studies and varies from 18.60% to 27.00%^38,40,41^, while the other four studies do not report gender distribution. Patients included were typically referred to CR, with one study specifying the prerequisite of being employed prior to a cardiac event^36^. The studies generally excluded patients lacking post-rehab data, those with contraindications to exercise, or those unwilling to participate. Functional sample sizes range significantly from 41 to 2280 across the studies.

**Table 1.** Characteristics of studies using ML prediction model in CR.

| Study (year) | Study location | Rehab phase | Study design | Age (years old) | Female (%) | Cohort inclusion criteria | Cohort exclusion criteria | Functional sample size |
| --- | --- | --- | --- | --- | --- | --- | --- | --- |
| Van et al (2010)<sup>35</sup> | Australia | II and III | Retrospective | Mean: 65.35 | NR | Patients referred to CR | Patients did not have post-rehab data or where values in target attributes were missing. | 2280 |
| Lofaro et al (2016)<sup>41</sup> | Italy | II | Retrospective | Mean: 67.98 | 18.60 | Patients admitted for CR after CAD or MI | NR | 129 |
| De Cannière et al (2020)<sup>38</sup> | Belgium | II | Retrospective | Mean: 63 | 27.00 | Patients eligible for CR | Patients who were unable to exercise or didn't complete the total study. | 89 |
| Jahandideh et al (2021)<sup>40</sup> | Australia | II | Cross-sectional | Mean: 63 | 26.27 | Patients referred to outpatient CR | Patients who were too ill or unable to speak English. | 217 |
| Yuan et al (2023)<sup>36</sup> | Malaysia | II | Retrospective | ≥18 | NR | Patients referred to CR and employed prior to having a cardiac event. | Patients who were pensioners or unemployed. | 184 |
| Filos et al (2023)<sup>37</sup> | Belgium and Ireland | III | Retrospective | 40-80 | NR | Patients completed a phase II CR and enrolled in Internet based home CR | Patients who dropped out or exercised very sparsely; | 41 |
| Torres et al (2023)<sup>39</sup> | Chile | II | Retrospective and prospective | ≥18 | NR | Patients referred to CR. | Patients who had any contraindication to physical exercise. | 227 |

Abbreviations: CAD: coronary artery disease

### Model characteristics

Table 2 presented a summary of characteristics of ML model characteristics included in our review. Each study has different prediction goals, ranging from forecasting a patient’s intention to begin CR to estimating their adherence to the program. Two studies aimed to predict patients’ prognosis in CR including 6-minute walk distance^38^, various physiological and psychological outcome^35^. Additionally, two studies focused on predicting patient disposition post-CR, such as likelihood of returning to work^36^ as well as the transition from one phase of CR to another^39^.

**Table 2.**
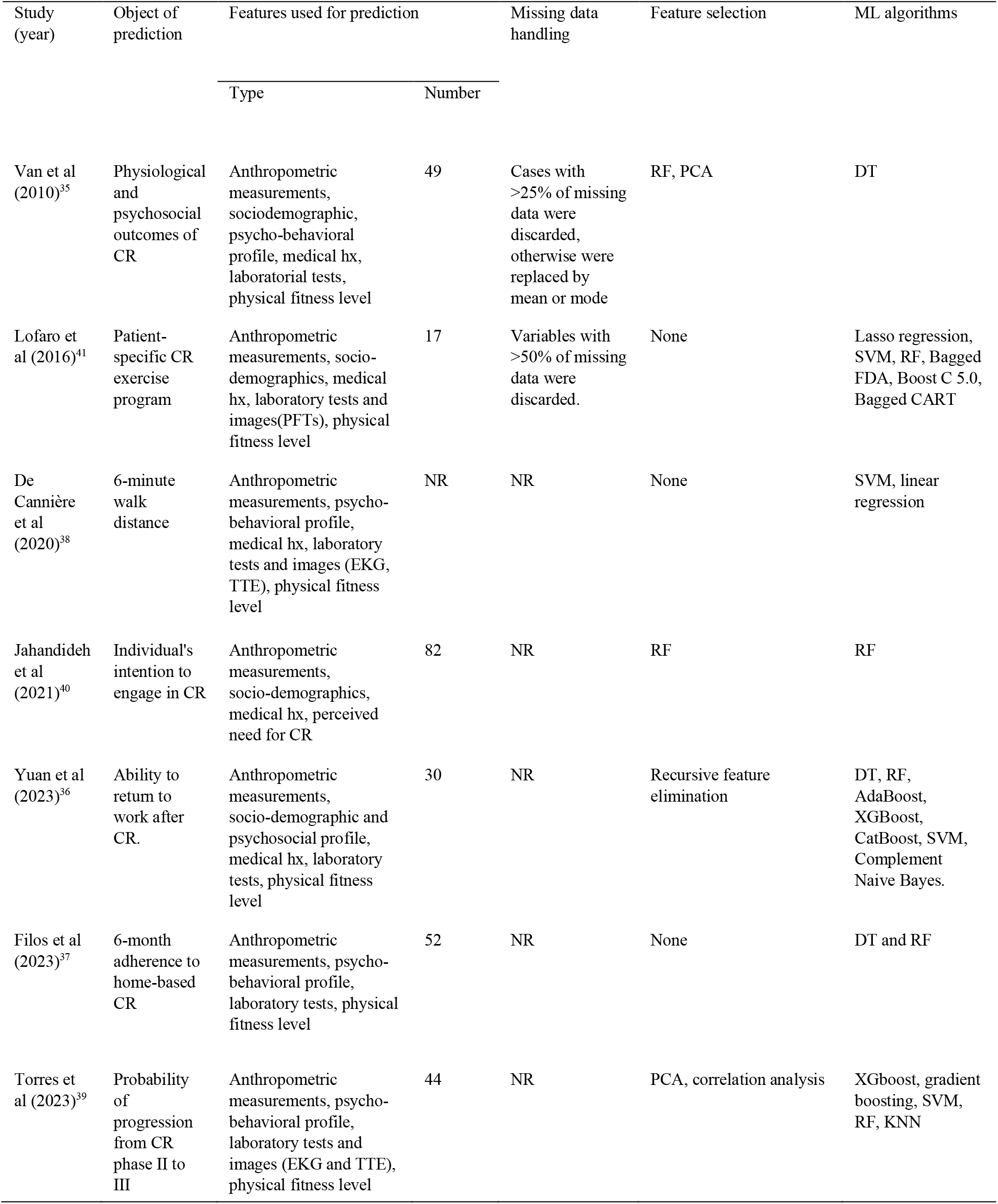
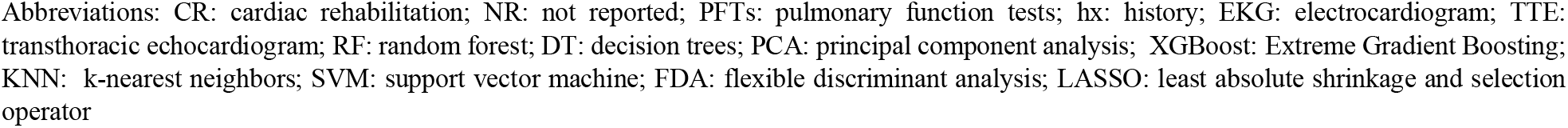
Characteristics of ML prediction model.

| Study (year) | Object of prediction | Features used for prediction |  | Missing data handling | Feature selection | ML algorithms |
| --- | --- | --- | --- | --- | --- | --- |
|  |  | Type | Number |  |  |  |
| Van et al (2010)<sup>35</sup> | Physiological and psychosocial outcomes of CR | Anthropometric measurements, sociodemographic, psycho-behavioral profile, medical hx, laboratorial tests, physical fitness level | 49 | Cases with >25% of missing data were discarded, otherwise were replaced by mean or mode | RF, PCA | DT |
| Lofaro et al (2016)<sup>41</sup> | Patient-specific CR exercise program | Anthropometric measurements, socio-demographics, medical hx, laboratory tests and images(PFTs), physical fitness level | 17 | Variables with >50% of missing data were discarded. | None | Lasso regression, SVM, RF, Bagged FDA, Boost C 5.0, Bagged CART |
| De Cannière et al (2020)<sup>38</sup> | 6-minute walk distance | Anthropometric measurements, psycho-behavioral profile, medical hx, laboratory tests and images (EKG, TTE), physical fitness level | NR | NR | None | SVM, linear regression |
| Jahandideh et al (2021)<sup>40</sup> | Individual's intention to engage in CR | Anthropometric measurements, socio-demographics, medical hx, perceived need for CR | 82 | NR | RF | RF |
| Yuan et al (2023)<sup>36</sup> | Ability to return to work after CR. | Anthropometric measurements, socio-demographic and psychosocial profile, medical hx, laboratory tests, physical fitness level | 30 | NR | Recursive feature elimination | DT, RF, AdaBoost, XGBoost, CatBoost, SVM, Complement Naive Bayes. |
| Filos et al (2023)<sup>37</sup> | 6-month adherence to home-based CR | Anthropometric measurements, psycho-behavioral profile, laboratory tests, physical fitness level | 52 | NR | None | DT and RF |
| Torres et al (2023)<sup>39</sup> | Probability of progression from CR phase II to III | Anthropometric measurements, psycho-behavioral profile, laboratory tests and images (EKG and TTE), physical fitness level | 44 | NR | PCA, correlation analysis | XGboost, gradient boosting, SVM, RF, KNN |

Abbreviations: CR: cardiac rehabilitation; NR: not reported; PFTs: pulmonary function tests; hx: history; EKG: electrocardiogram; TTE: transthoracic echocardiogram; RF: random forest; DT: decision trees; PCA: principal component analysis; XGBoost: Extreme Gradient Boosting; KNN: k-nearest neighbors; SVM: support vector machine; FDA: flexible discriminant analysis; LASSO: least absolute shrinkage and selection operator

These studies utilized a variety of patient features to develop their models. These features encompassed anthropometric measurements such as body mass index (BMI) and waist circumference, demographic information and medical history especially cardiovascular health. Psycho-behavioral profiles were also considered in most of the studies including evaluation of anxiety and depression levels. Laboratory test results and physical fitness levels were commonly used. Two studies harnessed specific imaging like EKG and TTE^38,39^.

The number of features used across the studies varies, with one utilizing as few as 17 features^41^ and others up to 82^40^. Most studies used pre-processing techniques for feature selection to reduce the number of input variables to those that are most important to the predictive model to improve performance and reduce computational cost. RF and principal component analysis (PCA) were most employed^35,39,40^.

Only two of the seven studies reported methods to handle missing data^35,41^. In terms of ML algorithms, there was a wide array employed across these studies, from basic decision trees (DT) and SVM to advanced ensemble methods like AdaBoost and XGBoost. Three of seven studies used a single algorithm^35,38,40^, the other four compared multiple different algorithms, with DT, RF and SVM being the most adopted^36,37,39,41^.

### Performance and validation

Table 3 outlined the best ML model with its performance metrics and validation approaches used to evaluate the ML models. The models’ performance was evaluated against various metrics such as area under the curve (AUC), sensitivity and specificity, mean absolute error (MAE), and normalized mean squared error (NMSE). The selection of best-performing ML algorithm differed per study. Six out of seven employed internal validation techniques, with cross-validation being the most prevalent method^35–39,41^. None of the studies underwent external validation, meaning they were not validated by using data from populations different from those used to develop the models, nor were they implemented in real-life practice.

**Table 3:** performance and validation of ML prediction model.

| Study (year) | Best ML algorithm | Performance metrics | Validation methods | External validation | Practice implementation |
| --- | --- | --- | --- | --- | --- |
| Van et al (2010)<sup>35</sup> | DT | AUC:0.815;<br>accuracy:0.769;<br>precision: 0.734;<br>sensitivity:0.769 | Internal validation:<br>10-fold cross<br>validation | None | None |
| Lofaro et al (2016)<sup>41</sup> | Lasso Regression | Accuracy: 0.935;<br>precision: 0.941;<br>sensitivity: 0.9 | 5-fold cross-validation; | None | None |
| De Cannière et al<br>(2020)<sup>38</sup> | SVM | MAE: 42.8 m | 20-fold-validation; | None | None |
| Jahandideh<br>et al (2021)<sup>40</sup> | RF | Accuracy: 0.715 | NR | None | None |
| Yuan et al (2023)<sup>36</sup> | AdaBoost | AUC:0.924;<br>accuracy: 0.864;<br>sensitivity: 0.928;<br>specificity: 0.733 | 10-fold cross<br>validation | None | None |
| Filos et al (2023)<sup>37</sup> | DT | Sensitivity:0.945;<br>precision: 0.80 | 10-fold cross<br>validation | None | None |
| Torres et al (2023)<sup>39</sup> | XGboost | NMSE: 0.008;<br>R<sup>2</sup>: 92%; | 10-fold cross<br>validation | None | None |

Abbreviations: DT: decision tree; RF: random forest; AUC: area under curve; SVM: support vector machine. MAE; mean absolute error (MAE);
NMSE: normalized mean squared error (NMSE); MAE: mean absolute error;

### Quality assessment

Table 4 summarized the scores for each dimension and the total score of each study according to the IMED checklist. The included studies had an average score of 30.8, with a range from 26 to 35.5. Most of the studies fell into the medium quality category, one study stood out as being of high quality^35^. Most studies demonstrated a discernible bias in the ‘Data Preparation’, ‘Validation’, and ‘Deployment’ dimensions, with lower scores that suggest these areas may have impacted the overall quality and reliability of the outcomes.

**Table 4.** Results of the quality assessment according to the IJMEDI checklist.

| Study(year) | Van et al<br>(2010)<sup>35</sup> | Lofaro et al<br>(2016)<sup>41</sup> | De<br>Cannière et<br>al (2020)<sup>38</sup> | Jahandideh<br>et al<br>(2021)<sup>40</sup> | Yuan et al<br>(2023)<sup>36</sup> | Filos et al<br>(2023)<sup>37</sup> | Torres et al<br>(2023)<sup>39</sup> |
| --- | --- | --- | --- | --- | --- | --- | --- |
| **Problem Understanding (10)** | 8.5 | 8.5 | 9.5 | 9 | 9.5 | 9.5 | 9 |
| 1 | 2 | 2 | 2 | 2 | 2 | 2 | 2 |
| 2 | 2 | 2 | 2 | 2 | 2 | 2 | 2 |
| 3 | 0.5 | 0.5 | 1 | 1 | 1 | 1 | 1 |
| 4 | 2 | 2 | 2 | 2 | 2 | 2 | 2 |
| 5 | 2 | 2 | 2 | 2 | 2 | 2 | 2 |
| 6 | 0 | 0 | 0.5 | 0 | 0 | 0 | 0 |
| **Data Understanding (6)** | 4 | 3 | 4 | 4 | 3 | 3 | 4 |
| 7 | 1 | 2 | 2 | 2 | 1 | 1 | 0 |
| 8 | 1 | 1 | 1 | 1 | 1 | 1 | 2 |
| 9 | 2 | 0 | 1 | 1 | 1 | 1 | 2 |
| **Data Preparation (8)** | 6 | 1 | 1 | 3 | 4 | 2 | 3 |
| 10 | 0 | 0 | 0 | 0 | 0 | 0 | 0 |
| 11 | 2 | 0 | 0 | 1 | 1 | 0 | 1 |
| 12 | 2 | 1 | 1 | 2 | 2 | 2 | 1 |
| 13 | 2 | 0 | 0 | 0 | 1 | 0 | 1 |
| **Modeling (6)** | 6 | 6 | 6 | 6 | 6 | 6 | 6 |
| 14 | 2 | 2 | 2 | 2 | 2 | 2 | 2 |
| 15 | 2 | 2 | 2 | 2 | 2 | 2 | 2 |
| 16 | 2 | 2 | 2 | 2 | 2 | 2 | 2 |
| **Validation (12)** | 8 | 6 | 6.5 | 4 | 7 | 6 | 8.5 |
| 17 | 2 | 1 | 1 | 1 | 2 | 2 | 2 |
| 18 | 2 | 2 | 2 | 1 | 1 | 1 | 2 |
| 19 | 0 | 0 | 0 | 0 | 0 | 0 | 0 |
| 20 | 2 | 1 | 1 | 1 | 2 | 2 | 2 |
| 21 | 0 | 0 | 0 | 0 | 0 | 0 | 0 |
| 22 | 2 | 2 | 2 | 1 | 2 | 1 | 2 |
| 23 | 0 | 0 | 0.5 | 0 | 0 | 0 | 0.5 |
| **Deployment (8)** | 2.5 | 1.5 | 3 | 3 | 3.5 | 2.5 | 3 |
| 24 | 1 | 1 | 1 | 1 | 1 | 1 | 1 |
| 25 | 0.5 | 0.5 | 0.5 | 1 | 1 | 0.5 | 0.5 |
| 26 | 0.5 | 0 | 1 | 0.5 | 1 | 0.5 | 1 |
| 27 | 0.5 | 0 | 0.5 | 0.5 | 0.5 | 0.5 | 0.5 |
| 28 | 0 | 0 | 0 | 0 | 0 | 0 | 0 |
| **29** | 0 | 0 | 0 | 0 | 0 | 0 | 0 |
| 30 | 0 | 0 | 0 | 0 | 0 | 0 | 0 |
| **Total (50)** | 35 | 26 | 30 | 30 | 33 | 29 | 33.5 |

\*Items in bold indicate high priority. 2, 1, 0 points for high-priority requirements that are, respectively, OK, mR and MR; assign half these scores (i.e., 1, 0.5 and 0) for low-priority requirement

## Discussion

This review has evaluated the performance of 22 ML based clinical models in 7 studies aiming to predict healthcare outcomes for patients participating in CR. The prediction objectives ranged from patient intention to initiate CR to graduate from outpatient CR along with interval physiological and psychological changes during the program. The best-performing ML models in their respective tasks reported AUC between 0.82 and 0.91, sensitivity from 0.77 to 0.95; demonstrating good prediction capabilities in general^42^. The majority of the included studies were rated as medium quality according to the IJMEDI checklist and there were high concerns for bias. Meta-analysis was not conducted as the included ML models were highly heterogenous in terms of targeted population, prediction objectives, outcome measurement and validation.

An ideal clinical prediction model should correctly distinguish between patients who will develop certain events and those who will not without misclassification in any case^33,43^. Its quality is associated with two properties of the model: discrimination and calibration. Discrimination is the model’s capacity to correctly separate individuals at higher risk of an event from those at lower risk. Calibration refers to the model’s ability to estimate absolute risks accurately^44^. Discrimination is typically measured by AUC of receiver operating characteristic (ROC) curve, it can also be assessed by sensitivity and specificity^45^. However, sensitivity and specificity vary as the cut point used to determine “positive” and “negative” test results change. The ROC curve is a graph of the sensitivity of a test versus its false-positive rate (1-specicificy) for all potential cut points. The AUC-ROC represents average prediction accuracy after balancing the inherent tradeoffs that exist between sensitivity and specificity across the spectrum of varying cut points^46^. A higher AUC indicates better discrimination ability. An AUC of 0.5 suggests no discrimination, equivalent to random guessing, while an AUC of 1.0 indicates perfect discrimination^47^. Van et al reported an AUC of 0.815, which suggests good discrimination in predicting post-rehab deterioration^35^. Yuan et al. reported an AUC of 0.923, reflecting the model’s excellent effectiveness in predicting patients’ likelihood of patients’ return-to-work^36^. Two studies reported sensitivity and precision rather than AUC as performance metrics, reflecting model’s performance for a particular cut point instead of all possible threshold^37,41^. One study reported accuracy only, it measures the proportion of true results, both true positives and true negatives. High accuracy can sometimes be misleading if the class distribution is uneven. Jahandideh et al. claimed an accuracy of 0.715 in differentiating highly motivated patients for CR initiation but did not report distribution of motivation levels in the study population^40^. This could lead to overestimation of accuracy in a predominantly motivated group or underestimation in a less motivated one. Overall, the reviewed studies used various evaluation metrics, appropriateness of which are generally acceptable, but could benefit from a more comprehensive measurement approach.

Model calibration is typically assessed after its discrimination is deemed acceptable. Calibration reflects the concordance between model prediction and observed outcomes. It is often represented by calibration plots or tables comparing predicted and observed risk ^44,48,49^. None of the included ML models underwent calibrations, so their ability to predict absolute risk remains uncertain. Although they have demonstrated overall good discrimination, calibration is essential to prove their capability in clinical decision support. This limitation contributed to lower scores in the deployment section of the IJMEDI checklist in table 4. Another major deficiency is the lack of external validation, which tests a model’s efficacy in a different population than it was initially derived from. Without robust external validation, a model’s generalizability is questionable ^48^. One notable example is that National Institute for Health and Care Excellence recommended an independent external validation of QRISK2 and the Framingham risk score, which were performed subsequently and demonstrated systematic miscalibration of Framingham risk scores and led to the need for different treatment thresholds in UK cohort^50^. Although some cases may not require immediate external validation if the sample size is large and representative of predictors and target outcomes on the top of appropriate internal validation^51^, most studies in our review, with populations ranging from 41 to 227 (except one with 2280 patients), would benefit from external validation as a key step towards implementation into clinical practice.

In addition to methodological robustness in model development, a useful clinical prediction model should address clinically significant issues. Low enrollment and adherence remain major challenges for CR^18^. Khatanga et al used logistic regression to identify factors such as surgical diagnosis, non or former tobacco use and intensity of physician recommendation as independent predictors for CR participation, whereas factors including anxiety, depression, or executive function had no significant impact^52^. Other studies have suggested that age, low socioeconomic status, gender, CR center location, psycho-behavioral factors including lack of motivation and reduced self-efficacy are barriers to participate in CR based on conventional statistics^53–55^. Our review included two studies which approached the issue via ML methods. Filos et al. developed an ML based prediction model to predict intention to engage in CR, incorporating all the aforementioned factors as predictors in model training. This model achieved a high sensitivity of 0.945. Additionally, the ML algorithm identified the contribution of each risk factor to the ultimate outcomes, offering potential guidance for providers on prioritizing clinical interventions in cases where multiple risk factors coexist^37^. Jahandideh et al predicted long term CR adherence using both identified risk factors mentioned above and physical fitness level, such as maximal quadriceps strength, which were not yet identified as independent risk factors. The model reached prediction accuracy at 71.5%. Some factors may be overlooked based on their individual statistical significance, but those factors may exert significant impact on ultimate outcome in combination with other factors. ML algorithms have the potential to identify complex interactions and patterns that traditional methods might miss^56,57^.

## Conclusion

There’s a scarcity of ML-based clinical prognostic models for predicting healthcare outcomes in CR patients. While current models show good prediction capacities, their missing information and methodological biases make it challenging to determine the best model or rank their performance. Future research should focus on developing new prediction models aiming at various outcomes in more diverse populations with robust methodological approaches. Additionally, enhancing existing models’ generalizability through external validation is needed. There’s a long journey ahead before these models can be fully embraced in clinical settings.

## Data Availability

All data produced in the present study are available upon reasonable request to the authors

## References

1. Tsao CW, Aday AW, Almarzooq ZI, et al. Heart Disease and Stroke Statistics—2023 Update: A Report From the American Heart Association. Circulation. 2023;147(8). doi:10.1161/CIR.0000000000001123

2. Heidenreich PA, Bozkurt B, Aguilar D, et al. 2022 AHA/ACC/HFSA guideline for the management of heart failure: a report of the American College of Cardiology/American Heart Association Joint Committee on Clinical Practice Guidelines. J Am Coll Cardiol. 2022;79(17):e263–e421.

3. Thomas RJ, Balady G, Banka G, et al. 2018 ACC/AHA Clinical Performance and Quality Measures for Cardiac Rehabilitation: A Report of the American College of Cardiology/American Heart Association Task Force on Performance Measures. Circ Cardiovasc Qual Outcomes. 2018;11(4). doi:10.1161/HCQ.0000000000000037

4. Lawton JS, Tamis-Holland JE, Bangalore S, et al. 2021 ACC/AHA/SCAI Guideline for Coronary Artery Revascularization: A Report of the American College of Cardiology/American Heart Association Joint Committee on Clinical Practice Guidelines. Circulation. 2022;145(3). doi:10.1161/CIR.0000000000001038

5. Virani SS, Newby LK, Arnold S V., et al. 2023 AHA/ACC/ACCP/ASPC/NLA/PCNA Guideline for the Management of Patients With Chronic Coronary Disease: A Report of the American Heart Association/American College of Cardiology Joint Committee on Clinical Practice Guidelines. Circulation. 2023;148(9). doi:10.1161/CIR.0000000000001168

6. Hamm LF, Sanderson BK, Ades PA, et al. Core Competencies for Cardiac Rehabilitation/Secondary Prevention Professionals. J Cardiopulm Rehabil Prev. 2011;31(1):2–10. doi:10.1097/HCR.0b013e318203999d

7. Anderson L, Thompson DR, Oldridge N, et al. Exercise-based cardiac rehabilitation for coronary heart disease. Cochrane Database of Systematic Reviews. Published online January 5, 2016. doi:10.1002/14651858.CD001800.pub3

8. Long L, Mordi IR, Bridges C, et al. Exercise-based cardiac rehabilitation for adults with heart failure. Cochrane Database of Systematic Reviews. 2019;2019(1). doi:10.1002/14651858.CD003331.pub5

9. Uithoven KE, Smith JR, Medina-Inojosa JR, Squires RW, Olson TP. The role of cardiac rehabilitation in reducing major adverse cardiac events in heart transplant patients. J Card Fail. 2020;26(8):645–651.

10. Francis T, Kabboul N, Rac V, et al. The Effect of Cardiac Rehabilitation on Health-Related Quality of Life in Patients With Coronary Artery Disease: A Meta-analysis. Canadian Journal of Cardiology. 2019;35(3):352–364. doi:10.1016/j.cjca.2018.11.013

11. Grochulska A, Glowinski S, Bryndal A. Cardiac Rehabilitation and Physical Performance in Patients after Myocardial Infarction: Preliminary Research. J Clin Med. 2021;10(11):2253. doi:10.3390/jcm10112253

12. Tessler J, Bordoni B. Cardiac Rehabilitation.; 2024.

13. Thomas RJ, Beatty AL, Beckie TM, et al. Home-based cardiac rehabilitation: a scientific statement from the American Association of Cardiovascular and Pulmonary Rehabilitation, the American Heart Association, and the American College of Cardiology. Circulation. 2019;140(1):e69–e89.

14. NCA - Cardiac Rehabilitation Programs (CAG-00089R) - Decision Memo. https://www.cms.gov/medicare-coverage-database/view/ncacal-decision-memo.aspx?proposed=N&NCAId=164.

15. Thomas RJ, Sapir O, Gomes PF, Iftikhar U, Smith JR, Squires RW. Advances, challenges, and progress in cardiac rehabilitation in chronic CVD management. Curr Atheroscler Rep. 2023;25(6):247–256.

16. Chindhy S, Taub PR, Lavie CJ, Shen J. Current challenges in cardiac rehabilitation: strategies to overcome social factors and attendance barriers. Expert Rev Cardiovasc Ther. 2020;18(11):777–789.

17. Ozemek C, Squires RW. Enrollment and Adherence to Early Outpatient and Maintenance Cardiac Rehabilitation Programs. J Cardiopulm Rehabil Prev. 2021;41(6):367–374. doi:10.1097/HCR.0000000000000645

18. Ritchey MD, Maresh S, McNeely J, et al. Tracking Cardiac Rehabilitation Participation and Completion Among Medicare Beneficiaries to Inform the Efforts of a National Initiative. Circ Cardiovasc Qual Outcomes. 2020;13(1). doi:10.1161/CIRCOUTCOMES.119.005902

19. Ngiam KY, Khor IW. Big data and machine learning algorithms for health-care delivery. Lancet Oncol. 2019;20(5):e262–e273. doi:10.1016/S1470-2045(19)30149-4

20. Rajula HSR, Verlato G, Manchia M, Antonucci N, Fanos V. Comparison of Conventional Statistical Methods with Machine Learning in Medicine: Diagnosis, Drug Development, and Treatment. Medicina (B Aires). 2020;56(9):455. doi:10.3390/medicina56090455

21. Azzolina D, Baldi (University of Padova) I, Barbati G, et al. Machine learning in clinical and epidemiological research: isn’t it time for biostatisticians to work on it? Epidemiol Biostat Public Health. 2022;16(4). doi:10.2427/13245

22. Haug CJ, Drazen JM. Artificial Intelligence and Machine Learning in Clinical Medicine, 2023. New England Journal of Medicine. 2023;388(13):1201–1208. doi:10.1056/NEJMra2302038

23. Ascent of machine learning in medicine. Nat Mater. 2019;18(5):407–407. doi:10.1038/s41563-019-0360-1

24. Muhammad LJ, Al-Shourbaji I, Haruna AA, Mohammed IA, Ahmad A, Jibrin MB. Machine learning predictive models for coronary artery disease. SN Comput Sci. 2021;2(5):350.

25. Dritsas E, Trigka M. Efficient data-driven machine learning models for cardiovascular diseases risk prediction. Sensors. 2023;23(3):1161.

26. Kanagarathinam K, Sankaran D, Manikandan R. Machine learning-based risk prediction model for cardiovascular disease using a hybrid dataset. Data Knowl Eng. 2022;140:102042.

27. Smith LA, Oakden-Rayner L, Bird A, et al. Machine learning and deep learning predictive models for long-term prognosis in patients with chronic obstructive pulmonary disease: a systematic review and meta-analysis. Lancet Digit Health. 2023;5(12):e872–e881. doi:10.1016/S2589-7500(23)00177-2

28. Zhang Z, Yang L, Han W, et al. Machine Learning Prediction Models for Gestational Diabetes Mellitus: Meta-analysis. J Med Internet Res. 2022;24(3):e26634. doi:10.2196/26634

29. Chowdhury MZI, Naeem I, Quan H, et al. Prediction of hypertension using traditional regression and machine learning models: A systematic review and meta-analysis. PLoS One. 2022;17(4):e0266334. doi:10.1371/journal.pone.0266334

30. Page MJ, Moher D, Bossuyt PM, et al. PRISMA 2020 explanation and elaboration: updated guidance and exemplars for reporting systematic reviews. bmj. 2021;372.

31. Riva JJ, Malik KMP, Burnie SJ, Endicott AR, Busse JW. What is your research question? An introduction to the PICOT format for clinicians. J Can Chiropr Assoc. 2012;56(3):167.

32. Cabitza F, Campagner A. The need to separate the wheat from the chaff in medical informatics: Introducing a comprehensive checklist for the (self)-assessment of medical AI studies. Int J Med Inform. 2021;153:104510.

33. Moons KGM, Wolff RF, Riley RD, et al. PROBAST: a tool to assess risk of bias and applicability of prediction model studies: explanation and elaboration. Ann Intern Med. 2019;170(1):W1–W33.

34. Wolff RF, Moons KGM, Riley RD, et al. PROBAST: a tool to assess the risk of bias and applicability of prediction model studies. Ann Intern Med. 2019;170(1):51–58.

35. Van A, Gay VC, Kennedy PJ, Barin E, Leijdekkers P. Understanding risk factors in cardiac rehabilitation patients with random forests and decision trees. In: Conferences in Research and Practice in Information Technology Series. ; 2010.

36. Choo JIA, Varathan KD, Suhaimi A, Lee WAN. Predicting Return to Work after Cardiac Rehabilitation using Machine Learning Models. J Rehabil Med. 2023;55.

37. Filos D, Claes J, Cornelissen V, Kouidi E, Chouvarda I. Predicting Adherence to Home-Based Cardiac Rehabilitation with Data-Driven Methods. Applied Sciences. 2023;13(10):6120.

38. De Cannière H, Corradi F, Smeets CJP, et al. Wearable monitoring and interpretable machine learning can objectively track progression in patients during cardiac rehabilitation. Sensors. 2020;20(12):3601.

39. Torres R, Zurita C, Mellado D, et al. Predicting Cardiovascular Rehabilitation of Patients with Coronary Artery Disease Using Transfer Feature Learning. Diagnostics. 2023;13(3):508.

40. Jahandideh S, Jahandideh M, Barzegari E. Individuals’ Intention to Engage in Outpatient Cardiac Rehabilitation Programs: Prediction Based on an Enhanced Model. J Clin Psychol Med Settings. Published online 2021:1-10.

41. Lofaro D, Groccia MC, Guido R, Conforti D, Caroleo S, Fragomeni G. Machine learning approaches for supporting patient-specific cardiac rehabilitation programs. In: 2016 Computing in Cardiology Conference (CinC). IEEE; 2016:149–152.

42. Li F, He H. Assessing the Accuracy of Diagnostic Tests. Shanghai Arch Psychiatry. 2018;30(3):207–212. doi:10.11919/j.issn.1002-0829.218052

43. Collins GS, Reitsma JB, Altman DG, Moons KGM. Transparent Reporting of a multivariable prediction model for Individual Prognosis Or Diagnosis (TRIPOD): The TRIPOD Statement. Ann Intern Med. 2015;162(1):55–63. doi:10.7326/M14-0697

44. Alba AC, Agoritsas T, Walsh M, et al. Discrimination and Calibration of Clinical Prediction Models. JAMA. 2017;318(14):1377. doi:10.1001/jama.2017.12126

45. Zou KH, O’Malley AJ, Mauri L. Receiver-Operating Characteristic Analysis for Evaluating Diagnostic Tests and Predictive Models. Circulation. 2007;115(5):654–657. doi:10.1161/CIRCULATIONAHA.105.594929

46. Obuchowski NA. Receiver operating characteristic curves and their use in radiology. Radiology. 2003;229(1):3–8.

47. Corbacioglu Ş, Aksel G. Receiver operating characteristic curve analysis in diagnostic accuracy studies: A guide to interpreting the area under the curve value. Turk J Emerg Med. 2023;23(4):195. doi:10.4103/tjem.tjem_182_23

48. Futoma J, Simons M, Panch T, Doshi-Velez F, Celi LA. The myth of generalisability in clinical research and machine learning in health care. Lancet Digit Health. 2020;2(9):e489–e492. doi:10.1016/S2589-7500(20)30186-2

49. Huang Y, Li W, Macheret F, Gabriel RA, Ohno-Machado L. A tutorial on calibration measurements and calibration models for clinical prediction models. Journal of the American Medical Informatics Association. 2020;27(4):621–633. doi:10.1093/jamia/ocz228

50. Collins GS, Altman DG. Predicting the 10 year risk of cardiovascular disease in the United Kingdom: independent and external validation of an updated version of QRISK2. Bmj. 2012;344.

51. Collins GS, Dhiman P, Ma J, et al. Evaluation of clinical prediction models (part 1): from development to external validation. BMJ. Published online January 8, 2024:e074819. doi:10.1136/bmj-2023-074819

52. Khadanga S, Savage PD, Gaalema DE, Ades PA. Predictors of Cardiac Rehabilitation Participation. J Cardiopulm Rehabil Prev. 2021;41(5):322–327. doi:10.1097/HCR.0000000000000573

53. Chindhy S, Taub PR, Lavie CJ, Shen J. Current challenges in cardiac rehabilitation: strategies to overcome social factors and attendance barriers. Expert Rev Cardiovasc Ther. 2020;18(11):777–789. doi:10.1080/14779072.2020.1816464

54. Oehler AC, Holmstrand EC, Zhou L, et al. Cost Analysis of Remote Cardiac Rehabilitation Compared With Facility-Based Cardiac Rehabilitation for Coronary Artery Disease. Am J Cardiol. 2024;210:266–272. doi:10.1016/j.amjcard.2023.08.061

55. Graversen CB, Eichhorst R, Ravn L, Christiansen SSR, Johansen MB, Larsen ML. Social inequality and barriers to cardiac rehabilitation in the rehab-North register. Scandinavian Cardiovascular Journal. 2017;51(6):316–322. doi:10.1080/14017431.2017.1385838

56. Badrulhisham F, Pogatzki-Zahn E, Segelcke D, Spisak T, Vollert J. Machine learning and artificial intelligence in neuroscience: A primer for researchers. Brain Behav Immun. 2024;115:470–479. doi:10.1016/j.bbi.2023.11.005

57. Sun GW, Shook TL, Kay GL. Inappropriate use of bivariable analysis to screen risk factors for use in multivariable analysis. J Clin Epidemiol. 1996;49(8):907–916. doi:10.1016/0895-4356(96)00025-x

