## Supplemental Table 1 for "Machine-Learning based Prediction Models for Healthcare Outcomes in Patients Participating in Cardiac Rehabilitation: A Systematic Review"

Supplementary table 1: searching strategy – from database inception to Jan 28, 2024

| Dataset | Google scholar |
| --- | --- |
| Searching strategy | "machine learning”OR“computational intelligence"OR"computer reasoning"OR"natural language process*"OR”random forest”OR"artificial intelligence"OR"unsupervised learning"OR"supervised learning"OR"deep learning"OR"neural network" AND "cardiac rehab*"OR”cardiovascular rehab*” |
| Results | 235 |
| Dataset | Pubmed |
| Searching strategy | Search: "cardiac rehabilitation" [mh] :3940  Search: Cardiac rehab*[tiab] OR cardiovascular rehab* [tiab] : 9180  Search: #1 or #2 : 10,155  Search: "Artificial Intelligence"[mh] :187680  Search: computational intelligence[tiab] OR machine intelligence[tiab] OR computer reasoning[tiab] OR computer vision[tiab] :9764  Search: random forest*[tiab] OR decision tree*[tiab] OR naive bayes*[tiab] OR support vector machine*[tiab] OR support vector network*[tiab] OR natural language process*[tiab] :69205  Search: artificial intelligence[tiab] OR machine learning[tiab] OR unsupervised learning[tiab] OR supervised learning[tiab] OR deep learning[tiab] OR neural network*[tiab] OR convolution*[tiab] OR residual network*[tiab] :253461  Search: #4 OR #5 OR #6 OR #7 :366983  Search: predict*[tiab] OR prognos*[tiab] OR prospective[tiab] : 3,307,547  Search: risk[tiab] AND (estimat*[tiab] OR score*[tiab] OR calculat*[tiab] OR index*[tiab] OR assess*[tiab] OR stratif*[tiab] OR validat*[tiab]) : 1,404,761  Search: c statistic[tiab] OR discriminat*[tiab] OR AUC[tiab] OR area under the curve[tiab] OR area under the receiver operator curve[tiab] OR risk ratio[tiab] OR odds ratio[tiab] OR hazard ratio[tiab] : 906,566  Search: #9 OR #10 OR #11 : 4,697,728  Search: #3 AND #8 AND #12 |
| Results | 23 |
| Dataset | Web of science |
| Searching strategy | #1 TS=("Cardiac rehab*" OR "Cardiovascular rehab*") :13290  #2 TS=("computational intelligence" or "machine intelligence" or "computer reasoning" or "computer vision") :85027  #3 TS=("random forest*" or "decision tree*" or "naive bayes*" or "support vector machine*" or "support vector network*" or "natural language process*") :271719  #4 TS=("artificial intelligence" or "machine learning" or "unsupervised learning" or "supervised learning" or "deep learning" or "neural network*" or "convolution*" or "residual network*") : 1218663  #5 #2 OR #3 OR #4 :1409700  #6 TS=("predict*" or "prognos*" or "prospective") :6011310  #7 TS=("risk" NEAR/5 ("estimat*" or "score*" or "calculat*" or "index*" or "assess*" or "stratif*" or "validat*")) :593590  #8 TS=("c statistic" or "discriminat*" or "AUC" or "area under the curve" or "area under the receiver operator curve" or "risk ratio" or "odds ratio" or "hazard ratio") :1186355  #9 #6 OR #7 OR #8 :7178933  #10 #1 AND # 5 AND #9 :22 |
| Results | 22 |
| Dataset | Scopus |
| Searching strategy | #1Title-Abs-Key( "Cardiac rehab*" OR "cardiovascular rehab*" ) :11869  #2 Title-Abs-Key("computational intelligence" or "machine intelligence" or "computer reasoning" or "computer vision") :234650  #3 Title-Abs-Key("random forest*" or "decision tree*" or "naive bayes*" or "support vector machine*" or "support vector network*" or "natural language process*") :459690  #4 Title-Abs-Key("artificial intelligence" or "machine learning" or "unsupervised learning" or "supervised learning" or "deep learning" or "neural network*" or "convolution*" or "residual network*") :2015641  #5 #2 OR #3 OR #4 :2395776  #6 Title-Abs-Key("predict*" or "prognos*" or "prospective") :7556306  #7 Title-Abs-Key("risk" W/5 ("estimat*" or "score*" or "calculat*" or "index*" or "assess*" or "stratif*" or "validat*")) :1345308  #8 Title-Abs-Key("c statistic" or "discriminat*" or "AUC" or "area under the curve" or "area under the receiver operator curve" or "risk ratio" or "odds ratio" or "hazard ratio") :1585611  #9 #6 OR #7 OR #8 :503579  #10 #1 AND #5 AND #9 :3 |
| Results | 3 |
